# Facemasks and similar barriers to prevent respiratory illness such as COVID-19: A rapid systematic review

**DOI:** 10.1101/2020.04.01.20049528

**Authors:** Julii Brainard, Natalia Jones, Iain Lake, Lee Hooper, Paul R Hunter

## Abstract

The current pandemic of COVID-19 has lead to conflicting opinions on whether wearing facemasks outside of health care facilities protects against the infection. To better understand the value of wearing facemasks we undertook a rapid systematic review of existing scientific evidence about development of respiratory illness, linked to use of facemasks in community settings.

**Methods:** We included all study designs. There were 31 eligible studies (including 12 RCTs). Narrative synthesis and random-effects meta-analysis of attack rates for primary and secondary prevention in 28 studies were performed. Results were reported by design, setting and type of face barrier in primary prevention, and by who wore the facemask (index patient or well contacts) in secondary prevention trials. The preferred outcome was influenza-like illness (ILI) but similar outcomes were pooled with ILI when ILI was unavailable. GRADE quality assessment was based on RCTs with support from observational studies.

**Results:** Where specific information was available, most studies reported about use of medical grade (surgical paper masks). In 3 RCTs, wearing a facemask may very slightly reduce the odds of developing ILI/respiratory symptoms, by around 6% (OR 0.94, 95% CI 0.75 to 1.19, I 29%, low-certainty evidence). Greater effectiveness was suggested by observational studies. When both house-mates and an infected household member wore facemasks the odds of further household members becoming ill may be modestly reduced by around 19% (OR 0.81, 95%CI 0.48 to 1.37, I 45%, 5 RCTs, low certainty evidence). The protective effect was very small if only the well person (OR 0.93, 95% CI 0.68 to 1.28, I 11%, 2 RCTs, low uncertainty evidence) or the infected person wore the facemask (very low certainty evidence).

**Discussion:** Based on the RCTs we would conclude that wearing facemasks can be very slightly protective against primary infection from casual community contact, and modestly protective against household infections when both infected and uninfected members wear facemasks. However, the RCTs often suffered from poor compliance and controls using facemasks. Across observational studies the evidence in favour of wearing facemasks was stronger. We expect RCTs to under-estimate the protective effect and observational studies to exaggerate it. The evidence is not sufficiently strong to support widespread use of facemasks as a protective measure against COVID-19. However, there is enough evidence to support the use of facemasks for short periods of time by particularly vulnerable individuals when in transient higher risk situations. Further high quality trials are needed to assess when wearing a facemask in the community is most likely to be protective.

## INTRODUCTION

On 30 January 2020 the World Health Organisation (WHO) declared a Public Health Emergency of International Concern (PHEIC) in response to the emergence of a novel coronavirus in Wuhan, China (World Health Organization, 2020a). In the declaration, the WHO stated there had been 170 deaths so far linked to the viral disease, later formally designated COVID-19. At the time of writing, more than 4600 deaths have been linked to COVID-19 (John Hopkins University Coronavirus Resource Centre, 2020) (accessed 12^th^ March 2020, 9:46am). On the 11^th^ March 2020 the WHO started referring to the COVID-19 epidemic as a pandemic (World Health Organization, 2020b). It is not clear when this outbreak will abate.

Amongst other advice widely sought by the public in response to this outbreak, was whether wearing face coverings, especially medical-grade coverings (e.g. masks, goggles or similar) might reduce the risk of catching or transmitting disease (Gajanan, 2020), particularly in domestic and public places. Despite no support by WHO encouraging wearing facemasks in the community, sales of inexpensive facemask products soared following the PHEIC declaration, leading to potential shortages for health care workers (Asgari and Wells, 2020; Carter, 2020; O’Connor, 2020; Taylor, 2020; Wu *et al*., 2020; US Surgeon General, 2020). Most previous similar systematic reviews investigating the effect of facemask use either focused solely on facemask use in health care settings or restricted study design to randomized controlled trials (RCTs), many of which were of low quality (MacIntyre and Chughtai, 2015).

We responded to this information demand by undertaking a rapid systematic review of the existing scientific evidence with regard to transmission of either respiratory disease or development of respiratory symptoms, especially influenza-like illness (ILI), linked to use of face barriers by non-health professionals and non-inpatients.

## METHODS

### Review question

Can wearing a face barrier (mask, goggles, shield, veil) in community settings prevent transmission of respiratory illness, such as from coronaviruses, rhinoviruses, influenza viruses or tuberculosis? We use the word ‘facemask’ as an umbrella term for diverse facial coverings that may cover any combination of mouth, nose and/or eyes.

### Search Strategy

Two recent literature reviews (MacIntyre and Chughtai, 2015; Jefferson *et al*., 2008) were consulted, to find eleven exemplar studies (Aiello *et al*., 2010; Aiello *et al*., 2012; Canini *et al*., 2010; Cowling *et al*., 2009; Cowling *et al*., 2008; Larson *et al*., 2010; Lau *et al*., 2004a; Lau *et al*., 2004b; Maclntyre *et al*., 2009; Simmerman *et al*., 2011; Suess *et al*., 2012) that met our eligibility criteria. We designed search strategies that were sensitive enough to find these exemplar studies and similar research, yet specific enough exclude most irrelevant records. The bibliographic databases Scopus, Embase and Medline were searched with the phrases in Box 1 on 31 Jan 2020. We also recent systematic reviews (Barasheed *et al*., 2016; Benkouiten *et al*., 2014; Cowling *et al*., 2010; MacIntyre and Chughtai, 2015; bin-Reza *et al*., 2012; Saunders-Hastings *et al*., 2017; Jefferson *et al*., 2008) on similar non-pharmaceutical practices for any missing primary studies.

### Assessment of inclusion

Two authors (JB and NJ) independently screened the retrieved titles and abstracts. Disagreements were resolved by discussion with other authors. The inclusion criteria were:

- Original research (not a review, guidelines, discussion, regulations, debate or commentary) published in English since January 1980
- The research needed to describe facemask use that might prevent disease transmission or symptom development among people in the community (rather than prevent transmission to or from clinically trained persons)
- The study described an observed relationship between facemask use and respiratory symptoms or infection by respiratory pathogens: (e.g. influenza, coronavirus, TB).
- There was a comparator or control group (non-barrier users, other barrier users or non-cases)
- Any study design in any country

The full text of each article that passed screening was retrieved and eligibility verified as part of data extraction.

**BOX 1 Bibliographic database search phrases**

##### SCOPUS

TITLE-ABS-KEY (

(facemask? OR “facemasks?” OR mask? OR goggle? OR faceshield? OR respirator OR respirators) AND

(influenza OR flu OR sars OR tuberculosis or mers OR coronav* OR “cov” OR respiratory-syndrome OR wuhan or “ncov”)

) AND

(LIMIT-TO (SUBJAREA, “MEDI”) OR LIMIT-TO (SUBJAREA, “NURS”) OR LIMIT-TO (SUBJAREA, “IMMU”))

##### EMBASE & Medline via the OVID portal

((facemask? or “face-masks?” or mask? or goggle? or face-shield? or respirator or respirators).ti.

or (facemask? or “face-masks?” or mask? or goggle? or face-shield? or respirator or respirators).ab.

or (facemask? or “face-masks?” or mask? or goggle? or face-shield? or respirator or respirators).kw.)

and

((influenza or flu or sars or tuberculosis or mers or coronav* or “cov” or respiratory-syndrome or “ncov” or wuhan).kw.

or

(influenza or flu or sars or tuberculosis or mers or coronav* or “cov” or respiratory-syndrome or “ncov” or wuhan).ti.

or

(influenza or flu or sars or tuberculosis or mers or coronav* or “cov” or respiratory-syndrome or “ncov” or wuhan).ab.)

### Data Extraction & risk of bias assessment

Qualitative data and numbers of participants who developed respiratory outcomes in relevant study arms were extracted. The preferred specific outcome was ILI, defined as fever ≥ 38 C° with cough and onset ≤ 10 days previous (World Health Organization, 2014). Where a WHO-definition was unavailable, we accepted other similar case definitions (e.g. cold symptoms, acute respiratory infections, clinical cases of influenza or SARS). Where studies reported three arms we extracted data for arms where the only difference was whether a facemask was worn (e.g. hand hygiene vs. hand hygiene + facemasks). Risk of bias in included RCTs was assessed (by LH) using the Cochrane risk of bias tool (Atkins *et al*., 2004), and biases and limitations identified by primary study authors of observational studies were noted. We assessed the quality of evidence using the GRADE framework, based on the RCT data and supported or contradicted by observational data (Atkins *et al*., 2004).

### Synthesis

All characteristics of included studies were tabulated. Numbers of infections and numbers of people at risk in each study arm were input to Review Manager 5.3 (The Cochrane Collaboration, 2014) for meta-analysis by JB, verified by NJ or LH. We calculated pooled odds ratios using Mantel*-*

Haenszel random effects meta-analysis (due to expected high heterogeneity) separately for primary prevention (when no cases were yet been identified) and prevention of secondary cases (when an individual was diagnosed with an infection and the aim was to prevent contacts from getting disease). We subgrouped by study design (RCT, pre/post, cohort, case-control or cross-sectional), and presented these subgroups in forest plots without global pooling to understand consistency of evidence across study designs. We also showed the trend of evidence within settings (subgrouping by setting). For secondary transmission (in RCTs) we subgrouped by who wore the facemask: index case, well contacts of the index case, or both. Outcomes after wearing faceveils were also presented where evidence was available.

A prospective patient (a mature adult living with many chronic health conditions including reduced lung function) commented on the draft manuscript and how the results could be made meaningful to laypeople.

## RESULTS

Altogether, 979 titles and abstracts were retrieved from Scopus, and 1443 from Embase and Medline combined. Our search located all 11 exemplar articles. Combining and deduplicating left 1725 articles. Of these 233 were not written in English and 81 were published before 1980, so were removed. This left 1458 titles and abstracts to screen, of which 46 were selected to be collected in full text. Full text review identified 26 eligible studies. Checking previous systematic reviews on protective effects of facemask use in the community identified a further five studies (all in the Hajj setting). Of the 31 included studies, 12 were cluster-RCTs, three were cohort studies, five were case-control and ten were cross-sectional, of which 28 reported data suitable for meta-analysis. Settings included schools, university residences, visits to health care providers, family households, the Hajj mass gathering, and non-specific community settings. Most studies reported on influenza-like illness (ILI) as an outcome (n=19), but fever with respiratory symptoms, upper respiratory tract infection, lab-confirmed or clinical influenza, toxic pneumonitis, common colds and other respiratory symptoms were used when ILI was unavailable. All mass gathering studies were associated with the Hajj pilgrimage. Table S1 lists the characteristics of the included studies.

### Prevention of primary infection

GRADE assessment suggested that wearing a facemask may very slightly reduce the odds of developing ILI/respiratory symptoms, by around 6% (OR 0.94, 95% CI 0.75 to 1.19, I^2^ 29%, low-certainty evidence, downgraded once each for risk of bias and imprecision). Figure 1 shows grouping of results by study design. Pooled data are presented to calculate a single odds ratio to compare and contrast study designs. Risk of biases for RCTs are also presented.

**Figure 1.**
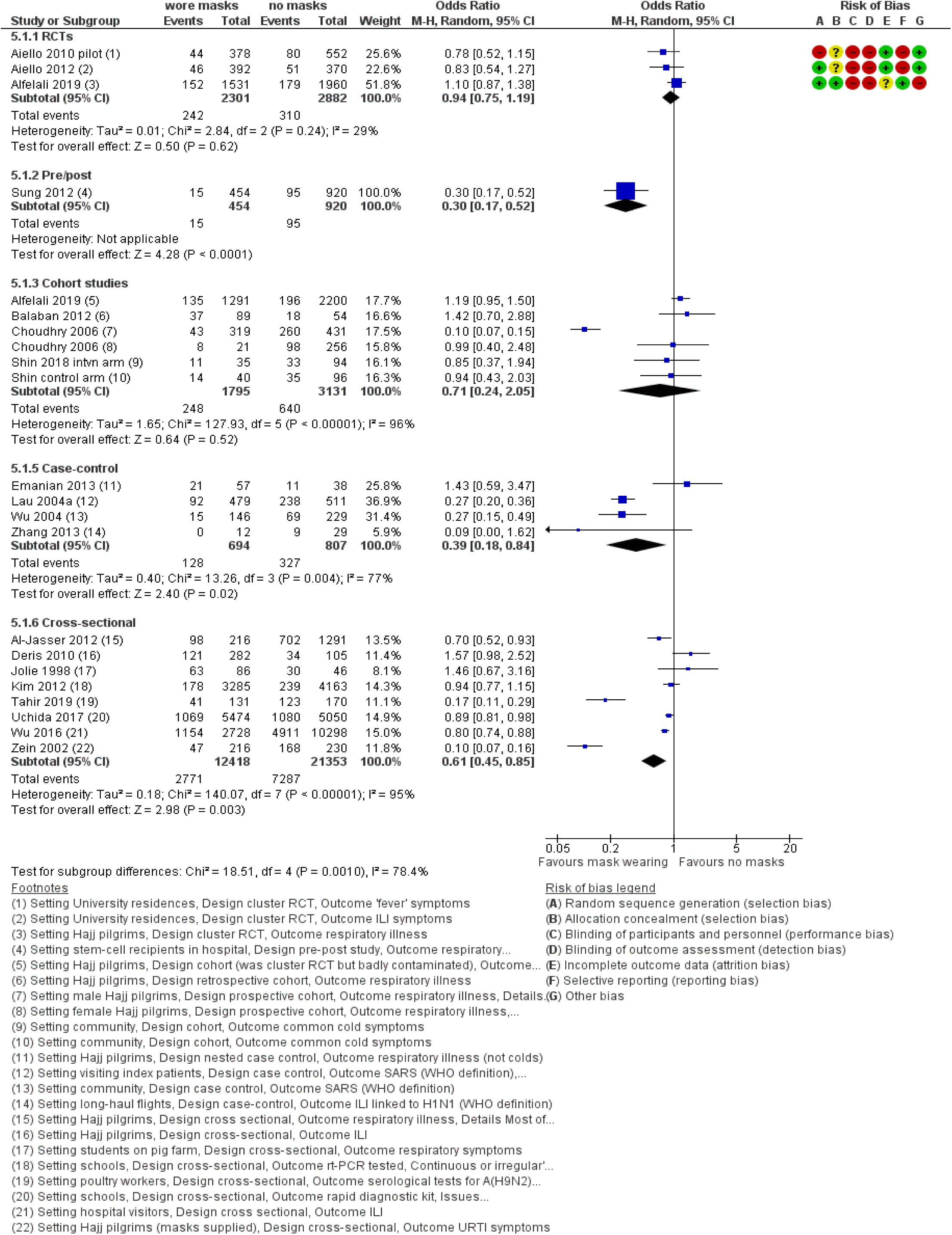
Mask wearing to prevent primary infection, by study design.

### Prevention of primary infection, subgrouping by study design

The three RCTs which measured the prevention of primary infection, indicated a slight, non-significant, reduction in the odds of primary infection with ILI (OR 0.94, 95%CI 0.75-1.19) (Figure 1, Comparison 1.1; Table 1). Heterogeneity was low (I^2^ 29%). Protection appeared greater in observational studies. Pre/post facemask wearing by visitors to hospital (Sung et al 2012) suggested a strong, significant reduction in infections among inpatient stem cell recipients (OR 0.30, 95%CI 0.17-0.52) (Figure 1, Comparison 1.2), but this finding is interpreted with caution because of likely changes in hygiene practices in the pre/post periods.

**Table 1.** Summary of GRADE findings

| Outcomes | Anticipated absolute effects*<br>(95% CI) |  | Relative effect<br>(95% CI) | No of<br>participants<br>(studies) | Certainty of the<br>evidence<br>(GRADE) | Comments |
| --- | --- | --- | --- | --- | --- | --- |
|  | Risk without<br>masks | Risk with<br>masks |  |  |  |  |
| Primary prevention, well wear masks - RCT data - outcome ILI | 108 per 1,000 | **102 per 1,000**<br>(83 to 125) | **OR 0.94**<br>(0.75 to 1.19) | 5183<br>(3 RCTs) | ⊕⊕○○<br>LOW<sup>a,b,c,d,e</sup> | Wearing a mask may very slightly reduce the odds of primary infection with influenza-like illness (ILI) by around 6%. Low-certainty evidence, downgraded once each for risk of bias and imprecision. |
| Secondary transmission, use of masks in homes, only ill person wears mask – RCT data – outcome ILI | 68 per 1,000 | **65 per 1,000**<br>(38 to 108) | **OR 0.95**<br>(0.53 to 1.72) | 903<br>(2 RCTs) | ⊕○○○<br>VERY LOW<sup>f,g</sup> | When one household member becomes ill with an ILI the effect of their wearing a mask on the odds of house-mates developing ILI is unclear, as the evidence is of very low certainty (downgraded once for risk of bias, twice for imprecision). |
| Secondary transmission, use of masks in homes, only well person(s) wears mask – RCT data – outcome ILI | 121 per 1,000 | **114 per 1,000**<br>(86 to 150) | **OR 0.93**<br>(0.68 to 1.28) | 2078<br>(2 RCTs) | ⊕⊕○○<br>LOW<sup>fh</sup> | Both house-mates and the infected household member wearing masks once one household member has contracted an infectious disease may modestly reduce the odds of further household members becoming ill by around 19%. Low certainty evidence (downgraded twice overall for risk of bias, imprecision and inconsistency). |
| Secondary transmission, use of masks in homes, both well and ill person(s) wear mask – RCT data – outcome ILI | 192 per 1,000 | **173 per 1,000**<br>(121 to 242) | **OR 0.81**<br>(0.48 to 1.37) | 1605<br>(5 RCTs) | ⊕⊕○○<br>LOW<sup>h,i,j</sup> | Both house-mates and the infected household member wearing masks once one household member has contracted an infectious disease may modestly reduce the odds of further household members becoming ill by around 19%. Low certainty evidence (downgraded twice overall for risk of bias, imprecision and inconsistency). |

\***The risk in the intervention group** (and its 95% confidence interval) is based on the assumed risk in the comparison group and the **relative effect** of the intervention (and its 95% CI). **CI**: Confidence interval; **OR**: Odds ratio

Evidence from the six cohort studies suggested facemasks provided some protection (OR 0.71, 95%CI 0.24-2.05), although, these findings were not significant (Figure 1, Comparison 1.3). Heterogeneity was very high (I = 96%) and the men-only cohort from Choudhry *et al*. (2006) was a noticeable outlier. This set of studies included observational data based on actual facemask wearing habits from one RCT (Alfelali *et al*. (2019)).

Among case-control (OR 0.39, 95%CI 0.18-0.84, I^2^ 77%) (Figure 1, Comparison 1.4) and cross-sectional studies (OR 0.61, 95%CI 0.45-0.85, I^2^ 95%) (Figure 1, Comparison 1.5), pooled data suggested that facemask wearing was protective, but effects were highly heterogeneous. Of the cross-sectional studies, Tahir *et al*. (2019) and Zein (2002) were noticeable outliers. Removal of these outliers still indicates facemask wearing as protective, although no longer significant, and heterogeneity falls slightly (OR 0.89, 95%CI 0.78-1.01, I = 64%) (data not shown).

Two further studies did not provide suitable data for pooling but reported that they had done analysis supporting their conclusions to comment narratively that facemasks were not protective (Hashim *et al*., 2016; Gautret *et al*., 2011). A third study without estimable data, Hashim *et al*. 2016 concluded that respirators were not effective protection against ILI.

### Prevention of primary infection by exposure setting

Figure 2 groups results by exposure setting. Pooling of data from different study designs is not appropriate to calculate a single odds ratio statistic. Most results favoured facemask wearing.

**Figure 2.**
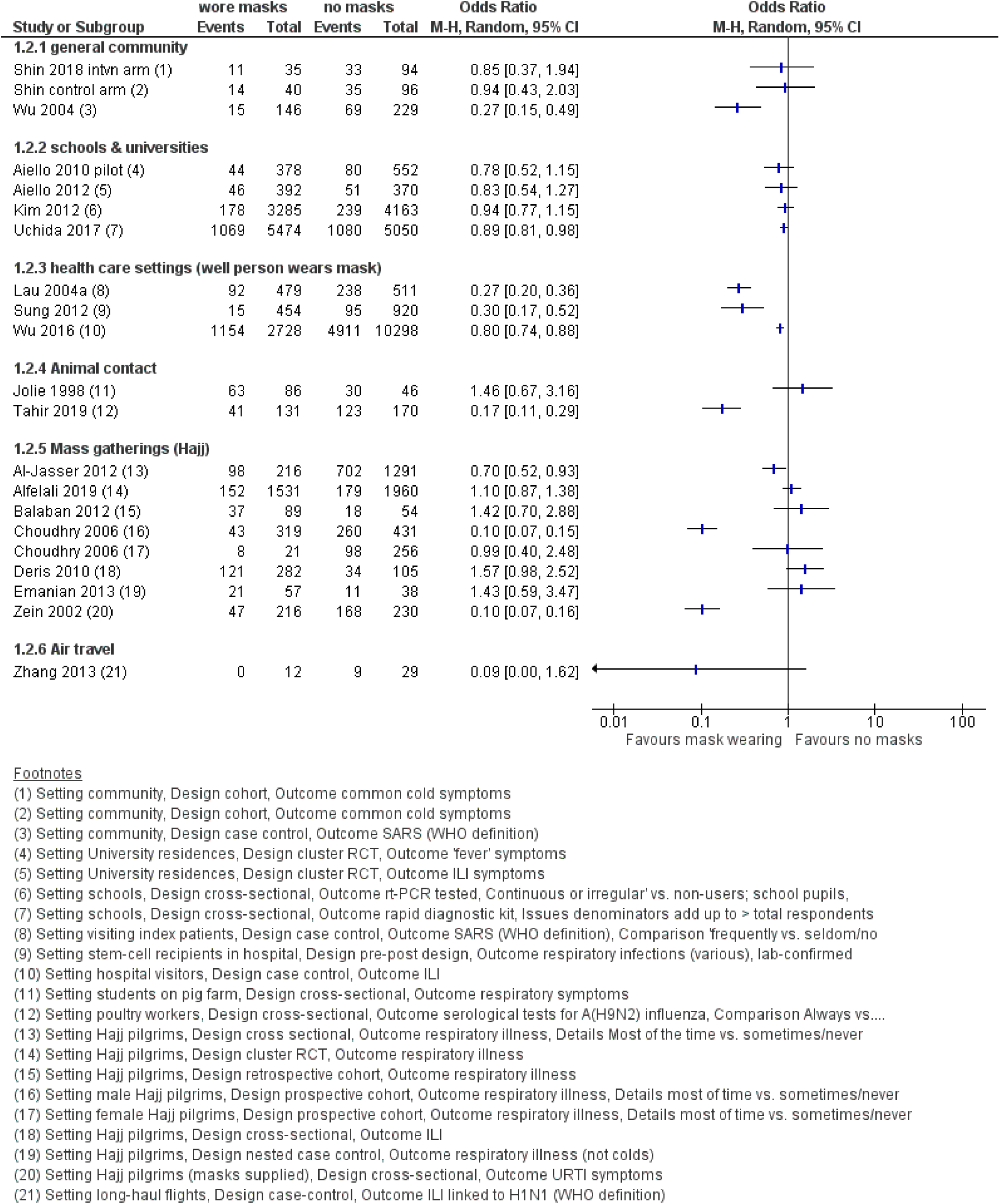
Mask wearing to prevent primary infection, by exposure setting.

Facemask wearing was consistently protective (the point-estimates of all included studies favoured facemask wearing) in the general community (2 cohort and 1 case-control; 1 significant protective finding) (Figure 2, Comparison 2.1), schools and universities (2 cluster-randomised RCTs and 2 cross-sectional; 1 significant protective finding) (Figure 2, Comparison 2.2), and for visits to health care clinics (1 pre/post and 2 case-control; 3 significant protective findings) (Figure 2, Comparison 2.3).

The results were less consistent (the point-estimates showed both protective and non-protective relationships) for animal contact (2 cross-sectional studies; 1 significant protective finding) (Figure 2, Comparison 2.4) and mass gatherings (all Hajj pilgrims; 1 cluster-randomised RCT, 2 cohort, 1 case-control and 3 cross-sectional; 3 significant protective findings) (Figure 2, Comparison 2.5).

One case-control study on air travel suggested a protective but non-significant relationship (Figure 2, Comparison 2.6).

### Prevention of primary infection among face veil wearers

Figure 3 shows data from two studies (cross-sectional and cohort) examining case incidence among women who wore face veils often/always while on Hajj pilgrimage. Both studies indicate a protective but non-significant relationship.

**Figure 3.**
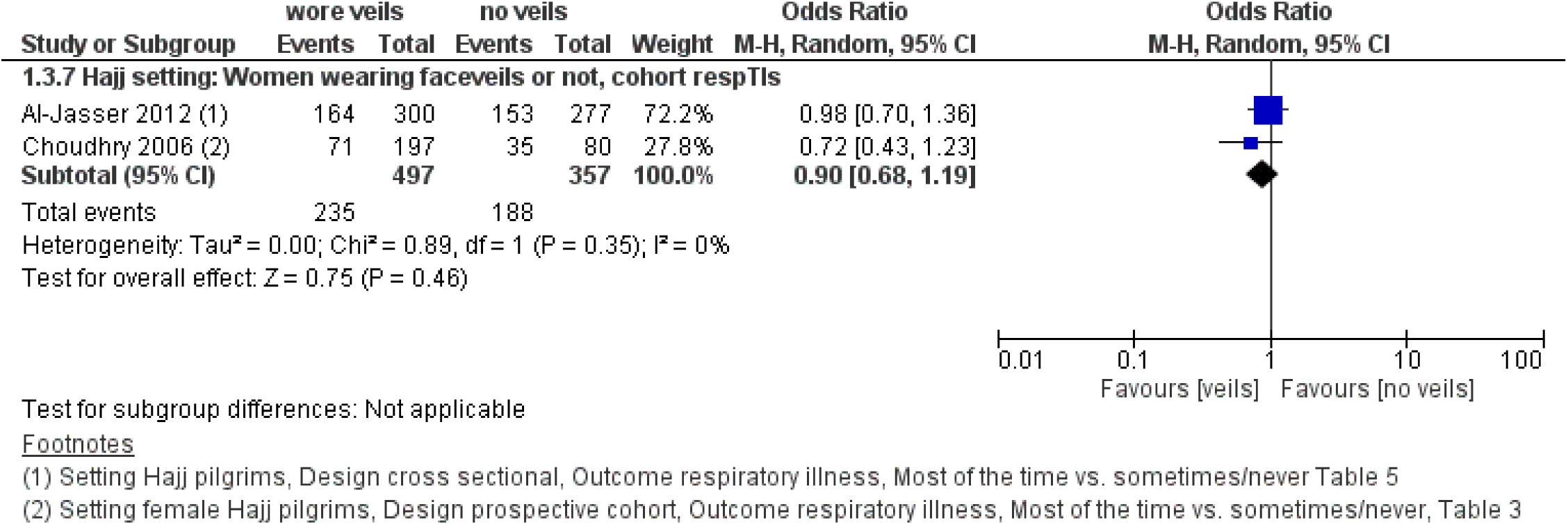
Faceveil wearing to prevent primary infection.

### Secondary transmission

GRADE assessment suggested that when both house-mates and an infected household member wore facemasks the odds of further household members becoming ill may be modestly reduced by around 19% (OR 0.81, 95%CI 0.48 to 1.37, I^2^ 45%, 5 RCTs, low certainty evidence, downgraded twice overall for risk of bias, imprecision and inconsistency). The protective effect was very small if only the well people wore facemasks (OR 0.93, 95% CI 0.68 to 1.28, I^2^ 11%, 2 RCTs, low uncertainty evidence, downgraded twice overall for risk of bias, imprecision and inconsistency) and unclear due to very low certainty evidence when only the infected person wore a facemask (downgraded once for risk of bias, twice for imprecision).

Figure 4 shows results for secondary transmission subdivided by study design and who wore the facemask (index patient, well contacts or both). Presented are pooled data to calculate a single odds ratio and risk of biases for RCTs. Findings from the two RCTs when only infected persons wore a facemask, suggested a very small, non-significant protective effect (OR 0.95, 95%CI 0.53 to 1.72, I^2^ 0%) (Figure 4, Comparison 4.1; Table 1). The evidence of a very small protective effect was stronger in the 2 RCTs where only well people wore facemasks (OR 0.93, 95%CI 0.68 to 1.28, I 11%) (Figure 4, Comparison 4.2; Table 1).

**Figure 4.**
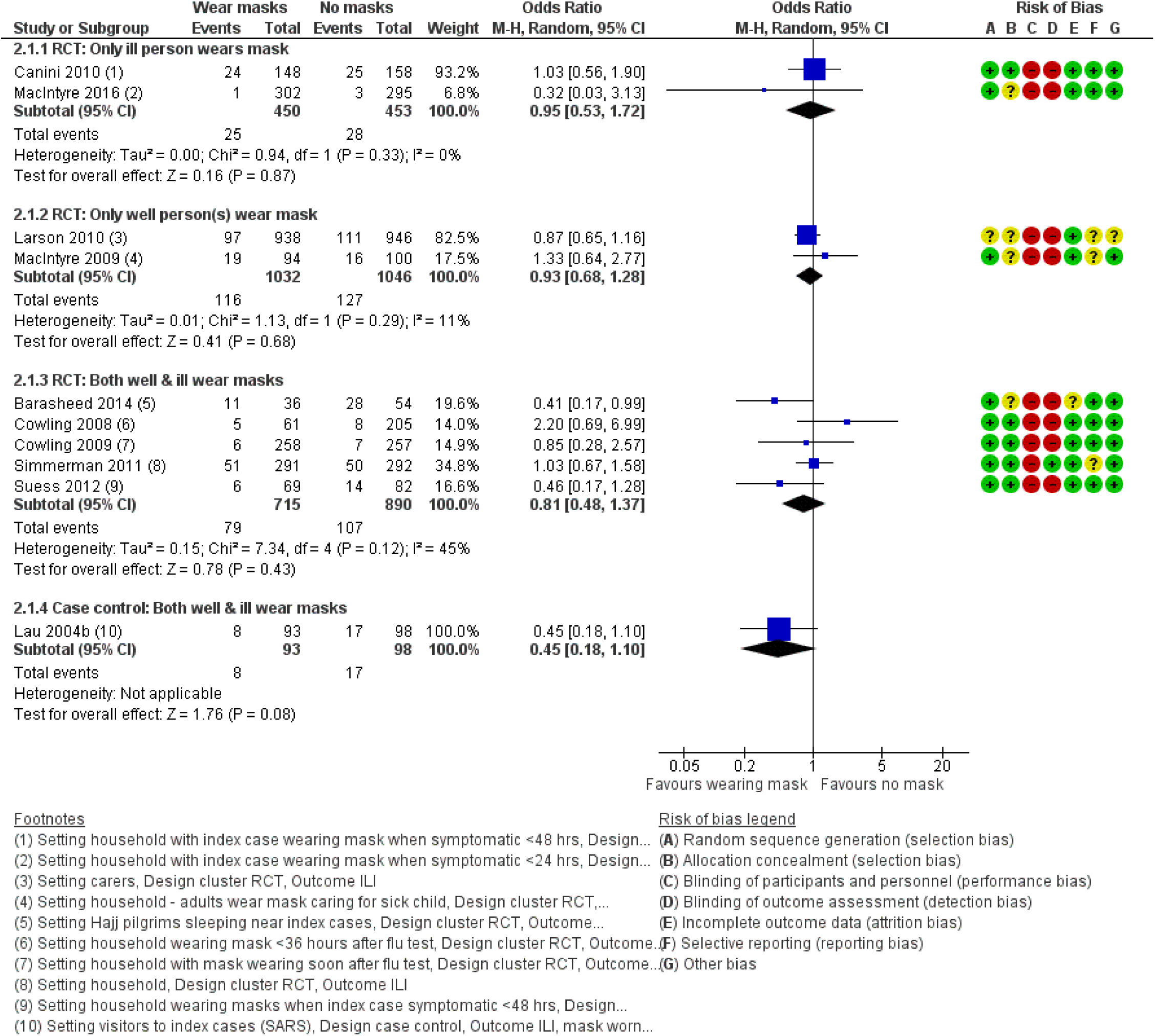
Mask wearing to prevent secondary infection, households.

Pooled data from five RCTs where both infected and non-infected household members wore facemasks showed the odds of infection fell modestly and not significantly (OR 0.81, 95%CI 0.48 to 1.37, I 45%) (Figure 4, Comparison 4.3, Table 1). Findings from the one case-control study where both infected and non-infected household members wore facemasks indicated a similar relationship (OR 0.45, 95%CI 0.18 to 1.10) (Figure 4, Comparison 4.4).

### Secondary transmission and commencement of facemask wearing

Figure 5 shows results for the four secondary transmission RCT studies providing data for attack rates when facemask wearing started < 36 hours after index patient became symptomatic. A single odds ratio statistic and risk of biases for RCTs are presented.

**Figure 5.**
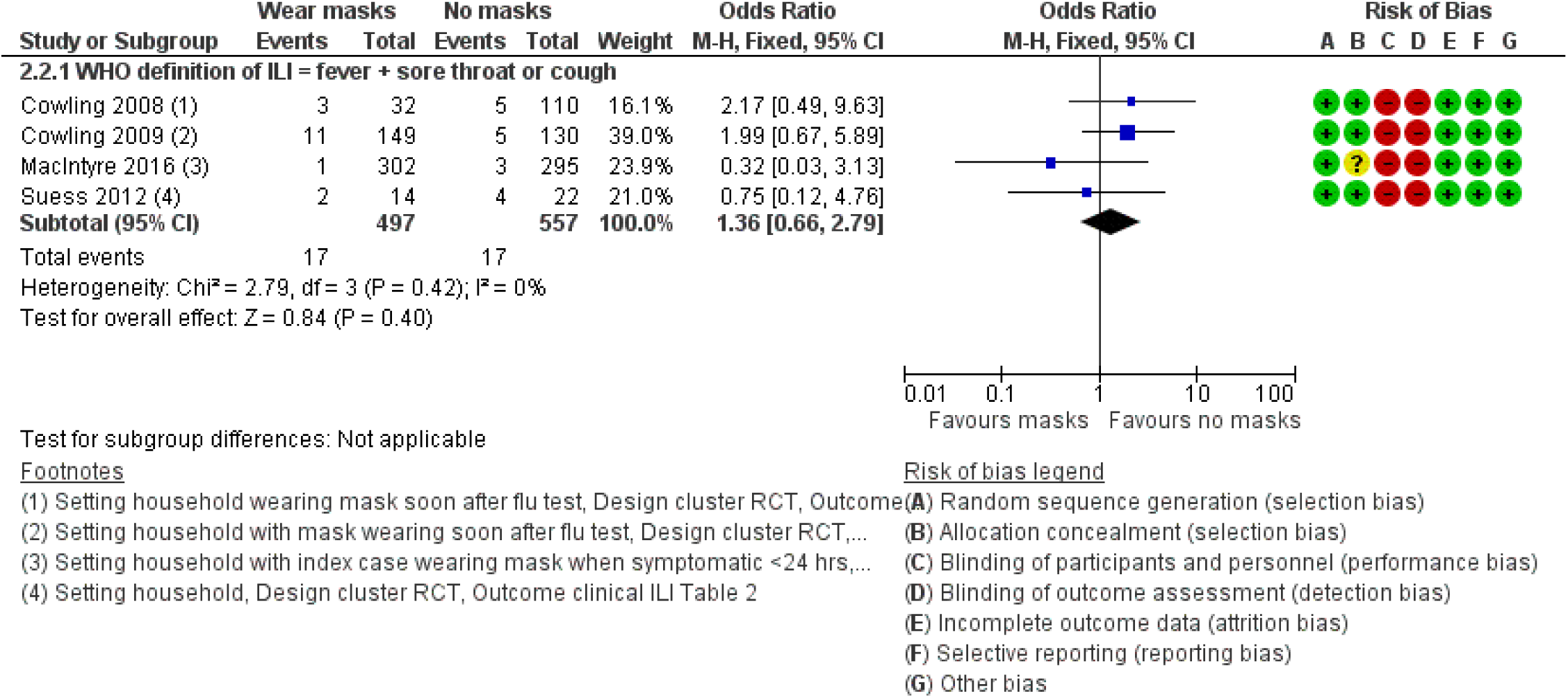
Mask wearing to prevent secondary infection starting <36 hours after onset in index patient, households.

Facemask wearing was not protective in this subgroup analysis OR 1.36, 0.66-2.79, I^2^ 0%). Some of the original investigators in these studies undertook logistic regression to adjust their findings for other confounders and found evidence that early facemask wearing (< 36 hours after symptom onset) could be protective, but acknowledged that their models were underpowered.

## DISCUSSION

In this study we have undertaken a rapid review to examine whether the use of facemasks impacts on the transmission of respiratory symptoms. Evidence based on RCTs suggests that wearing a facemask may very slightly reduce the odds of primary infection with ILI by around 6% (low-certainty evidence). Observational studies suggested greater effectiveness. In households where infected people were grouped with uninfected people, if both house-mates and the infected household member wore facemasks the odds of further household members becoming ill were reduced by around 19% (low certainty evidence). Where only the uninfected people wore facemasks the effect was very small (reducing the odds of infection by 7%, low certainty evidence). The evidence is similar where only the infected household members wore facemasks (reducing the odds of infection by 5%, very low certainty evidence). However, due to controls wearing facemasks when they shouldn’t and poor compliance (intervention participants not wearing facemasks when they should) we expect that RCT evidence under-estimated efficacy. In contrast, we expect observational studies, especially those based on self-reported symptoms, have over-estimated how protective facemask wearing can be because of confounding. Therefore, specific accurate estimates of the degree of protectiveness of facemasks are not feasible from the currently available evidence base.

Facemasks appear be most effective when worn to prevent primary respiratory illness in relatively low risk situations: community settings where contact may be casual and relatively brief, such as on public transport, in shops, in workplaces and perhaps in university residences or schools with limited shared public spaces. Facemask wearing is probably not protective during mass gatherings, but evidence on use during mass gatherings is inconsistent. All studies focussed on Hajj pilgrimage which may not be a typical mass gathering event (especially large and prolonged). Facemask wearing within households where infection was already present was modestly effective in the included studies, and this evidence was fairly consistent (low-medium heterogeneity, I from 0% to 45%). Limited effectiveness of primary prevention at Hajj or secondary prevention within households may be because of the multiplicity of transmission pathways within these settings and high level of recurring contact. It may also be due to the late use of facemasks, usually > 24 hours after a household or group member became symptomatic which could be 48 hours after they became infectious (Centers for Disease Control, 2018).

That facemasks might protect wearers has been cast in doubt during the COVID-19 outbreak (eg., Abramson, 2020; Geggel, 2020; Harris, 2020), often supported by the observation that surgical facemasks were designed (originally) to protect patients from the wearers, and that facemasks soon become very moist with condensation from wearer’s breath (facilitating microbial ingress and growth). Nevertheless, worn correctly for brief periods, wearing surgical masks have been shown to provide an average 6-fold reduction in exposure to aerosolized influenza virus (Booth *et al*., 2013), so it is unsurprising that facemask wearing was linked to fewer cases in our synthesis, especially primary observational studies in community settings. With regard to secondary attack rates, whether the ill or the initially well people in a household wore facemasks (or both, or neither wore facemasks) did not seem to matter: attack rates were similar in household-setting RCTs. These details suggest that other factors (not who wore the facemask, but rather duration and type of contact) also matter greatly to disease transmission. Many barriers exist that can make it difficult for individuals to wear facemasks correctly for hours over a multi-day period, including perceived breathing impairment and other discomforts (Sim *et al*., 2014; Suess *et al*., 2011). Facemasks are perceived to or genuinely do interfere with ordinary physical activities such as heavy exertion, sleep, oral hygiene and eating. Facemasks can be uncomfortable, hot, cause skin rashes or simply feel anti-social (Al Badri, 2017; Donovan *et al*., 2007).

Previous systematic literature reviews on the efficacy of using facemasks in community settings are often combined with other personal protection measures (Saunders-Hastings *et al*., 2017; Jefferson *et al*., 2008; Wong *et al*., 2014) or mixed health care workers with non-health care workers (Jefferson *et al*., 2008; bin-Reza *et al*., 2012; Wang *et al*., 2015). Those that have looked specifically at community use have focused only on RCTs (Cowling *et al*., 2010; MacIntyre and Chughtai, 2015). Overall, the reviews had mixed conclusions about community settings: that facemasks were highly effective (Jefferson *et al*., 2008), definitely effective (Barasheed *et al*., 2016; bin-Reza *et al*., 2012), may be effective for protection (Benkouiten *et al*., 2014; Cowling *et al*., 2010; MacIntyre and Chughtai, 2015) or had a not statistically significant protective effect (Saunders-Hastings *et al*., 2017). There was near consensus that the evidence base was inadequate (Cowling *et al*., 2010; bin-Reza *et al*., 2012; Barasheed *et al*., 2016; MacIntyre and Chughtai, 2015; Saunders-Hastings *et al*., 2017; Benkouiten *et al*., 2014).

RCTs may not be the best quality evidence to evaluate a population behaviour like facemask use that is likely to be imperfectly implemented. Many of the included RCTs reported that participants did not follow instructions (Alfelali *et al*., 2019; Barasheed *et al*., 2016; Cowling *et al*., 2009; Cowling *et al*., 2008; Maclntyre *et al*., 2009; MacIntyre *et al*., 2016). In these RCTs, large proportions of controls wore facemasks during the monitoring period (Simmerman *et al*., 2011; MacIntyre *et al*., 2016; Cowling *et al*., 2008), while intervention participants did not wear facemasks the majority of the time (Maclntyre *et al*., 2009; MacIntyre *et al*., 2016; Larson *et al*., 2010; Cowling *et al*., 2009; Cowling *et al*., 2008). Compared to RCTs, observational data (cohort and case-control studies) may give superior quality evidence for efficacy of facemask wearing to avoid ILI, given they are trying to relate actual behaviour to outcomes. Of course, observational data are subject to recall bias and other confounding factors that RCTs may obviate. For expediency, we have not undertaken formal quality assessment on the diverse observational studies included in this review: we expect that the observational evidence would be found to have many potential sources of bias that RCT designs are intended to avoid. We found original study authors gave many informative comments on their own study limitations (Supplemental Table 1). We argue that there is merit in combining study types to generate a full view of existing evidence.

A difficulty with previous reviews and also in our own review, is the mix of facial barrier used. All of the RCTs included in our review provided specific facemasks (usually surgical grade, rarely P2 or equivalent grade respirator) with instructions how to wear the facemask, as well as told users the frequency facemasks should be changed and how to hygienically dispose of used facemasks. Information was not reported about the types of facemasks that (contrary to protocol) some controls in RCTs used. Only a few of the observational studies collected specific information about what type of face covering was used.

We assume that where not explicitly stated, most facemasks worn were surgical grade in the included observational studies. This view concords with survey of facemasks visible in recent street scene photography of Hong Kong, Korean, Chinese and Japanese cities where facemask wearing is common: most of the evident facemasks resemble surgical styles and is somewhat supported by surveys and discussions of habitual facemask users, usually in Far East countries (Wada *et al*., 2012; Shirai, 2019; Hung, 2018; Burgess and Horii, 2012).

Future investigations should collect information about what facemasks people had, how the facemasks were worn, whether they were reused and how they were prepared for reuse, as well as duration of facemask wear. The impact of the timing of intervention is worth analysing further especially if studies can be fully powered to produce more definitive results, or if evidence should emerge about facemask use within homes before symptom onset or within a very short period (perhaps 4-12 hours) after symptom onset.

## Limitations

The search strategy tried to include all ‘respiratory’ illnesses. However, actual transmission risks vary with the type of pathogen: adequate protection for one germ may be unsafe against others. All of the articles we found related to viruses, most typically influenza but also SARS coronavirus. The lack of tuberculosis studies included in this review may relate to the date cut off we imposed (we omitted studies published before 1980 for expediency). If the evidence base were larger then we could meaningfully subset the studies by the types of pathogen studied: e.g. coronaviruses vs. influenza. Similarly, although we preferred clinical case definitions of ILI or respiratory illness (based on symptoms), where only lab-confirmed case definitions were available, we grouped these with clinical cases. The case definitions were also based mostly on self-report. These pathogens, outcomes and reporting origins could be meaningfully separated with a larger evidence base.

Due to the rapidity of this review we did not consider other article archives or databases such as Google Scholar, CINAHL and MedRXiv. We did not undertake quality assessment of observational studies, although we have provided the authors own assessments of their study limitations which highlight many potential issues. We did not classify outcomes by severity of symptoms or other clinical outcomes (Paulo *et al*., 2010). It is possible that facemask wearing reduced duration or severity of symptoms experienced due to reducing infectious dose received, although not actual disease. Severity or duration of symptoms and other specific clinical outcomes are important to patients and this is an area which requires further research. We also didn’t separate results by duration that facemasks were worn; some studies did subgroup analysis that found this influenced attack rates.

The only mass-gathering setting where facemask wearing evidence has been gathered and published is the Hajj. This event may not be comparable to many smaller mass-gatherings and the Hajj is exceptional for high contact levels (Zein, 2002; Deris *et al*., 2010; Shirah *et al*., 2017; Hashim *et al*., 2016; Gautret *et al*., 2011).

We have not undertaken a full cost-benefit analysis. The sudden emergence of COVID-19 led to high community demand for face barriers and raised valid concerns that insufficient supplies of facemasks were available for health care workers (Wu *et al*., 2020; US Surgeon General, 2020). The environmental and economic costs of regularly using facemasks are notable, and only partly abated by reuse. Wearing facemasks consistently and correctly for long periods may not be easy for many people. There is a need to calculate where the balance of benefits and costs lie in facemask wearing for disease prevention (Baracco *et al*., 2015; Rivera *et al*., 1997; McGain *et al*., 2017; Coulter, 2017).

We make no comment on the relative utility of other proposed protective measures compared to facemask wearing, such as self-isolation or frequent handwashing: we have not undertaken research on those measures for comparison. We did not formally assess likelihood of publication bias in the primary research evidence base, due to this being a rapid review.

## CONCLUSIONS

Our study was a rapid review of the literature on the impact of the use of facemasks on the transmission of respiratory symptoms. Based on randomized trial data, wearing a facemask may very slightly reduce the odds of primary infection with influenza-like illness (ILI). However, included trials generally had very low compliance and comparisons undermined by some controls also wearing facemasks. Considering both experimental and observational studies, we consider that the balance of evidence is that facemasks may offer some protection against primary respiratory virus infection when used in community settings outside the home. We surmise that this relates to relatively short exposure windows, brief duration of wear and casual contact, while noting that existing evidence is prone to recall bias and lack of information about what kind of facemasks were worn or if they were worn correctly. Facemasks appear to be slightly effective for preventing secondary infection within household settings, but the certainty of this evidence is low. Facemasks used in the Hajj-related multi-day mass gatherings (which typically involve 2-6 weeks of inter-related activities) have not consistently prevented respiratory symptoms from developing. We do not consider that the balance of evidence across all available studies supports routine and widespread use of facemasks in the community. However, using a mask for short periods of time by particularly vulnerable individuals during transient exposure events may be justified.

## Data Availability

REVMAN file that holds the data available upon request.

**SUPPLEMENTAL** See Table S1 for full list of included studies.

## ACKNOWLEDGEMENTS

Thanks to Dalal Ardan for explaining what it’s like to be a Hajj pilgrim. Alison Aiello kindly and swiftly sent supplemental information for her research. A mature adult member of the public with chronic health conditions (including sarcoidosis) provided helpful comments on how to make the article meaningful to lay persons.

## Notes

### Competing Interest Statement

The authors have declared no competing interest.

### Funding Statement

No funder supported this study.

## REFERENCES

Abramson, A. 2020. Surgical Masks Won’t Protect You Against the Coronavirus. Allure.

Aiello, A. E., Murray, G. F., Perez, V., Coulborn, R. M., Davis, B. M., Uddin, M., Shay, D. K., Waterman, S. H. & Monto, A. S. 2010. Mask use, hand hygiene, and seasonal influenza-like illness among young adults: A randomized intervention trial. Journal of Infectious Diseases, 201, 491–498.

Aiello, A. E., Perez, V., Coulborn, R. M., Davis, B. M., Uddin, M. & Monto, A. S. 2012. Facemasks, hand hygiene, and influenza among young adults: A randomized intervention trial. PLoS ONE, 7, e29744.

Al Badri, F. M. 2017. Surgical mask contact dermatitis and epidemiology of contact dermatitis in healthcare workers. Current Allergy & Clinical Immunology, 30, 183–188.

Alfelali, M., Haworth, E. A., Barasheed, O., Badahdah, A.-M., Bokhary, H., Tashani, M., Azeem, M. I., Kok, J., Taylor, J. & Barnes, E. H. 2019. Facemask versus No Facemask in Preventing Viral Respiratory Infections During Hajj: A Cluster Randomised Open Label Trial. SSRN (Lancet preprints).

Asgari, N. & Wells, P. 2020. Face mask shortage hits Europe and US as coronavirus spreads. Financial Times, 30 Jan.

Atkins, D., Best, D., Briss, P. & Et Al 2004. GRADE Working Group. Grading quality of evidence and strength of recommendations. BMJ, 328.

Baracco, G., Eisert, S., Eagan, A. & Radonovich, L. 2015. Comparative cost of stockpiling various types of respiratory protective devices to protect the health care workforce during an influenza pandemic. Disaster Medicine and Public Health Preparedness, 9, 313–318.

Barasheed, O., Alfelali, M., Mushta, S., Bokhary, H., Alshehri, J., Attar, A. A., Booy, R. & Rashid, H. 2016. Uptake and effectiveness of facemask against respiratory infections at mass gatherings: A systematic review. International Journal of Infectious Diseases, 47, 105–111.

Benkouiten, S., Brouqui, P. & Gautret, P. 2014. Non-pharmaceutical interventions for the prevention of respiratory tract infections during Hajj pilgrimage. Travel Medicine and Infectious Disease, 12, 429–442.

Bin-Reza, F., Lopez Chavarrias, V., Nicoll, A. & Chamberland, M. E. 2012. The use of masks and respirators to prevent transmission of influenza: A systematic review of the scientific evidence. Influenza and other Respiratory Viruses, 6, 257–267.

Booth, C. M., Clayton, M., Crook, B. & Gawn, J. M. 2013. Effectiveness of surgical masks against influenza bioaerosols. Journal of Hospital Infection, 84, 22–26.

Burgess, A. & Horii, M. 2012. Risk, ritual and health responsibilisation: Japan’s ‘safety blanket’of surgical face mask-wearing. Sociology of Health & Illness, 34, 1184–1198.

Canini, L., Andreoletti, L., Ferrari, P., Angelo, D. R., Blanchon, T., Lemaitre, M., Filleul, L., Ferry, J. P., Desmaizieres, M., Smadja, S., Valleron, A. J. & Carrat, F. 2010. Surgical mask to prevent influenza transmission in households: A cluster randomized trial. PLoS One, 5.

Carter, S. L. 2020. Sold-Out Coronavirus N95 Face Masks Offer a Lesson in Price Gouging. Bloomberg.com, 31 Jan.

Centers for Disease Control. 2018. How Flu Spreads [Online]. Available: https://www.cdc.gov/flu/about/disease/spread.htm [Accessed 17 Feb 2020].

Choudhry, A., Al Mudaimegh, K., Turkistani, A. & Al Hamdan, N. 2006. Hajj-associated acute respiratory infection among hajjis from Riyadh. Eastern Mediterranean Health Journal, 12, 300–309.

Coulter, J. 2017. Air pollution masks symptom of throwaway society. China Daily, 18 Jan.

Cowling, B. J., Chan, K. H., Fang, V. J., Cheng, C. K. Y., Fung, R. O. P., Wai, W., Sin, J., Seto, W. H., Yung, R., Chu, D. W. S., Chiu, B. C. F., Lee, P. W. Y., Chiu, M. C., Lee, H. C., Uyeki, T. M., Houck, P. M., Peiris, J. S. M. & Leung, G. M. 2009. Facemasks and hand hygiene to prevent influenza transmission in households: A cluster randomized trial. Annals of Internal Medicine, 151, 437–446.

Cowling, B. J., Fung, R. O. P., Cheng, C. K. Y., Fang, V. J., Chan, K. H., Seto, W. H., Yung, R., Chiu, B., Lee, P., Uyeki, T. M., Houck, P. M., Peiris, J. S. M. & Leung, G. M. 2008. Preliminary findings of a randomized trial of non-pharmaceutical interventions to prevent influenza transmission in households. PLoS ONE, 3, e2101.

Cowling, B. J., Zhou, Y., Ip, D. K. M., Leung, G. M. & Aiello, A. E. 2010. Face masks to prevent transmission of influenza virus: a systematic review. Epidemiology and Infection, 138, 449–456.

Deris, Z. Z., Hasan, H., Sulaiman, S. A., Wahab, M. S. A., Naing, N. N. & Othman, N. H. 2010. The prevalence of acute respiratory symptoms and role of protective measures among Malaysian Hajj pilgrims. Journal of Travel Medicine, 17, 82–88.

Donovan, J., Kudla, I., Holness, L. D., Skotnicki-Grant, S. & Nethercott, J. R. 2007. Skin reactions following use of N95 facial masks. Dermatitis, 18, 104.

Gajanan, M. 2020. Can Face Masks Prevent Coronavirus? Experts Say That Depends. Time.

Gautret, P., Vu Hai, V., Sani, S., Doutchi, M., Parola, P. & Brouqui, P. 2011. Protective measures against acute respiratory symptoms in French pilgrims participating in the Hajj of 2009. Journal of Travel Medicine, 18, 53–55.

Geggel, L. 2020. Can wearing a face mask protect you from the new coronavirus? Live Science.

Harris, D. M. 2020. Spokesperson for World Health Organisation. In: Ferrari, N. (ed.) Breakfast is Leading Britain’s Conversation. LBC Radio.

Hashim, S., Ayub, Z. N., Mohamed, Z., Hasan, H., Harun, A., Ismail, N., Rahman, Z. A., Suraiya, S., Naing, N. N. & Aziz, A. A. 2016. The prevalence and preventive measures of the respiratory illness among malaysian pilgrims in 2013 hajj season. Journal of Travel Medicine, 23.

Hung, Y.-W. 2018. A Study of Barriers to the Wearing of Face Masks by Adults in the US to Prevent the Spread of Influenza. Masters of Science in Design, Arizona State University.

Jefferson, T., Foxlee, R., Del Mar, C., Dooley, L., Ferroni, E., Hewak, B., Prabhala, A., Nair, S. & Rivetti, A. 2008. Physical interventions to interrupt or reduce the spread of respiratory viruses: Systematic review. BMJ, 336, 77–80.

John Hopkins University Coronavirus Resource Centre. 2020. Coronavirus COVID-19 Global Cases by the Center for Systems Science and Engineering (CSSE) at Johns Hopkins University (JHU) [Online]. JHU. Available: https://coronavirus.jhu.edu/map.html [Accessed 12th March, 9:46am 2020].

Larson, E. L., Ferng, Y. H., Wong-Mcloughlin, J., Wang, S., Haber, M. & Morse, S. S. 2010. Impact of non-pharmaceutical interventions on URIs and influenza in crowded, urban households. Public Health Reports, 125, 178–191.

Lau, J. T. F., Lau, M., Kim, J. H., Wong, E., Tsui, H. Y., Tsang, T. & Wong, T. W. 2004a. Probable Secondary Infections in Households of SARS Patients in Hong Kong, Emerging Infectious Diseases. 10 (2) (pp 235–243), 2004. Date of Publication: February 2004.

Lau, J. T. F., Tsui, H., Lau, M. & Yang, X. 2004b. SARS Transmission, Risk Factors, and Prevention in Hong Kong, Emerging Infectious Diseases. 10 (4) (pp 587–592), 2004. Date of Publication: April 2004.

Macintyre, C. R. & Chughtai, A. A. 2015. Facemasks for the prevention of infection in healthcare and community settings. BMJ (Online), 350.

Macintyre, C. R., Zhang, Y., Chughtai, A. A., Seale, H., Zhang, D., Chu, Y., Zhang, H., Rahman, B. & Wang, Q. 2016. Cluster randomised controlled trial to examine medical mask use as source control for people with respiratory illness. BMJ Open, 6.

Maclntyre, C. R., Cauchemez, S., Dwyer, D. E., Seale, H., Cheung, P., Browne, G., Fasher, M., Wood, J., Gao, Z., Booy, R. & Ferguson, N. 2009. Face mask use and control of respiratory virus transmission in households. Emerging Infectious Diseases, 15, 233–241.

Mcgain, F., Story, D., Lim, T. & Mcalister, S. 2017. Financial and environmental costs of reusable and single-use anaesthetic equipment. British Journal of Anaesthesia, 118, 862–869.

O’connor, K. 2020. Coronavirus: face masks sell out but are unlikely to stop germs. The Times, 1 Feb.

Paulo, A. C., Correia-Neves, M., Domingos, T., Murta, A. G. & Pedrosa, J. 2010. Influenza infectious dose may explain the high mortality of the second and third wave of 1918–1919 influenza pandemic. PLoS One, 5.

Rivera, P., Louther, J., Mohr, J., Campbell, A., Dehovitz, J. & Sepkowitz, K. A. 1997. Does a cheaper mask save money? The cost of implementing a respiratory personal protective equipment program. Infection Control and Hospital Epidemiology, 18, 24–27.

Saunders-Hastings, P., Crispo, J. a. G., Sikora, L. & Krewski, D. 2017. Effectiveness of personal protective measures in reducing pandemic influenza transmission: A systematic review and meta-analysis. Epidemics, 20, 1–20.

Shirah, B. H., Zafar, S. H., Alferaidi, O. A. & Sabir, A. M. 2017. Mass gathering medicine (Hajj Pilgrimage in Saudi Arabia): The clinical pattern of pneumonia among pilgrims during Hajj. Journal of Infection and Public Health, 10, 277–286.

Shirai, N. 2019. Japanese Citizens Becoming Masked Heroes to Serve Society. BA, Princeton.

Sim, S. W., Moey, K. S. P. & Tan, N. C. 2014. The use of facemasks to prevent respiratory infection: A literature review in the context of the Health Belief Model. Singapore Medical Journal, 55, 160–167.

Simmerman, J. M., Suntarattiwong, P., Levy, J., Jarman, R. G., Kaewchana, S., Gibbons, R. V., Cowling, B. J., Sanasuttipun, W., Maloney, S. A., Uyeki, T. M., Kamimoto, L. & Chotipitayasunondh, T. 2011. Findings from a household randomized controlled trial of hand washing and face masks to reduce influenza transmission in Bangkok, Thailand. Influenza and other Respiratory Viruses, 5, 256–267.

Suess, T., Remschmidt, C., Schink, S., Luchtenberg, M., Haas, W., Krause, G. & Buchholz, U. 2011. Facemasks and intensified hand hygiene in a German household trial during the 2009/2010 influenza A(H1N1) pandemic: Adherence and tolerability in children and adults. Epidemiology and Infection, 139, 1895–1901.

Suess, T., Remschmidt, C., Schink, S. B., Schweiger, B., Nitsche, A., Schroeder, K., Doellinger, J., Milde, J., Haas, W., Koehler, I., Krause, G. & Buchholz, U. 2012. The role of facemasks and hand hygiene in the prevention of influenza transmission in households: Results from a cluster randomised trial; Berlin, Germany, 2009-2011. BMC Infectious Diseases, 12.

Tahir, M. F., Abbas, M. A., Ghafoor, T., Dil, S., Shahid, M. A., Bullo, M. M. H., Ain, Q. U., Ranjha, M. A., Khan, M. A. & Naseem, M. T. 2019. Seroprevalence and risk factors of avian influenza H9 virus among poultry professionals in Rawalpindi, Pakistan. Journal of Infection and Public Health, 12, 482–485.

Taylor, K. 2020. Costco is selling out of surgical masks in South Korea, as the country battles the spread of the coronavirus. Business Insider, 3 Feb.

The Cochrane Collaboration 2014. Review Manager (RevMan). 5.3 ed. Copenhagen: The Nordic Cochrane Centre.

Us Surgeon General 2020. Seriously people - STOP BUYING MASKS! Coronavirus Disease 19 (COVID-19). Twitter.

Wada, K., Oka-Ezoe, K. & Smith, D. R. 2012. Wearing face masks in public during the influenza season may reflect other positive hygiene practices in Japan. BMC Public Health, 12.

Wang, M., Barasheed, O., Rashid, H., Booy, R., El Bashir, H., Haworth, E., Ridda, I., Holmes, E. C., Dwyer, D. E., Nguyen-Van-Tam, J., Memish, Z. A. & Heron, L. 2015. A cluster-randomised controlled trial to test the efficacy of facemasks in preventing respiratory viral infection among Hajj pilgrims. Journal of Epidemiology and Global Health, 5, 181–189.

Wong, V. W. Y., Cowling, B. J. & Aiello, A. E. 2014. Hand hygiene and risk of influenza virus infections in the community: A systematic review and meta-analysis. Epidemiology and Infection, 142, 922–932.

World Health Organization. 2014. WHO surveillance case definitions for ILI and SARI [Online]. Available: https://www.who.int/influenza/surveillance_monitoring/ili_sari_surveillance_case_definition/en/ [Accessed 14 Feb 2020].

World Health Organization. 2020a. Statement on the second meeting of the International Health Regulations (2005) Emergency Committee regarding the outbreak of novel coronavirus (2019-nCoV) [Online]. WHO. Available: https://www.who.int/news-room/detail/30-01-2020-statement-on-the-second-meeting-of-the-international-health-regulations-(2005)-emergency-committee-regarding-the-outbreak-of-novel-coronavirus-(2019-ncov) [Accessed 7 Feb 2020].

World Health Organization. 2020b. WHO Director-General’s opening remarks at the media briefing on COVID-19 - 11 March 2020 [Online]. WHO. Available: https://www.who.int/dg/speeches/detail/who-director-general-s-opening-remarks-at-the-media-briefing-on-covid-1911-march-2020 [Accessed 12th March 2020].

Wu, H., Huang, J., Zhang, C. J., He, Z. & Ming, W.-K. 2020. Facemask shortage and the novel coronavirus (2019-nCoV) outbreak: Reflection on public health measures. medRxiv.

Zein, U. The role of using masks to reduce acute upper respiratory tract infections in pilgrims. 4th Asia Pacific travel health conference, Oct 20 2002 Shanghai, PR China.

